# Regional mortality variations among older adults in India: Evidence from Demographic Health Survey

**DOI:** 10.1101/2022.05.22.22275427

**Authors:** Saddaf Naaz Akhtar, Nandita Saikia

## Abstract

**Background:** Studies on regional mortality variations among older adults in India are poorly documented. Therefore, we intend to estimate the impact of individual and district level determinants on regional mortality variations among the older adults in India. Additionally, we have performed decomposition analysis to evaluate the contribution of socioeconomic and demographic factors in the inter-regional mortality variations among older adults in India.

**Methods:** We performed a two-level logistic regression model using data from Demographic Health Survey (2015-16) for India to quantify the impact of socio-demographic and ecomonic characteristics. We have also analyzed multivariate decomposition approach to determine the role of determinants in regional mortality variations among older adults.

**Results:** The multilevel analyses results revealed that regional mortality variations exist at both individual and district levels among older adults in India. Our finding identified that older adults living in the Central region have a higher mortality risk than in Northern regions. The decomposition results showed that the Central, Eastern and North-eastern regions have significantly higher average number of excess mortality compared to other regions among older adults. The district-level literacy, insurance coverage, electricity supply and public health facilities also showed a significant impact on district level mortality among older adults in India.

**Conclusion:** Our study insights extremely important determinants for India’s public health. In order to eliminate these mortality gaps, there is a need for solid support from the state and central government to bridge the socio-demographic and economic development in India at the regional level. As a result, policy should include efforts to improve health outcomes among older adults at early stages.

## 1. Introduction

Old age mortality still remains a major health concern and challenge in most developing nations. Old age mortality is demographically defined as deaths of individual age 60 or 65 years and above. It is closely connected with the outcome of the key developments goals such as the Millennium Development Goals (MDGs) which are now transformed to Sustainable Development Goals (SDGs) (United Nations, 2021). Improvement in health such as combating of non-communicable diseases and other diseases are the part of this goals where old age people play a prominent role. Therfore, a significant impact on morbidity and mortality among the older adults affects the outcomes of these goals as well as the progress of a country or nation socially, demographically, and economically.

Though mortality among older adults is declining in most countries, however the they seem to be gaps in the pace of decline and period stagnation but some converges between European countries, (Janssen et al., 2003, 2004; Mesle & Vallin, 2006; Vierboom et al., 2019; Wilson et al., 2020), contributing to population ageing, which still remains a health burden for many developing countries. Globally, 5.6 million older adults died in 2019, and the death rate for India is accounted for the people 60+ is 42.6 deaths per 1000 where the male is 45.9 deaths per 1000 and female is 39.5 deaths per 1000 in 2018 (Office of the Registrar General & Census Commissioner, India, 2018). These differences also exist in both rural and urban settings or areas. On the other hand, old age mortality is a serious public health issue with wide-ranging socioeconomic ramifications for households, communities, and the country. There is a dearth of studies that produce research on old age mortality at the regional level in India. Furthermore, the majority of the previous studies have paid much attention to infant (Dyson & Moore, 1983; Gupta et al., 2016; Jain, 1985), child (Bora, 2020; Liu et al., 2019) and adult mortality (Panda & Mishra, 2021; Saikia et al., 2011; Saikia & Moradhvaj, 2021) than old age mortality; as a result, more is known about regional variations in infant, child and adult mortality than the latter age in India. Therefore, our study attempted to fill this gap.

Studying regional mortality variations is crucial as it is an essential indicator of public health through which population can be forecast. It helps in the local planning and implementing health policy in the society at the regional settings. Ageing population in regional areas face many challenges, hence studying regional mortality variations would help in increasing health care needs and provide financial support and other services. Accessibility of adequate health services may get affected and eventually resulting to regional variations in mortality and health. Therefore, it is important to shed light into the role of changing regional population compositions that provides important information for the health professional and policymakers.

Regional mortality variations among the older adults have not been well documented yet in India. However, the question arises, which socioeconomic, demographic and district level predictors of mortality can be identified and how great are resulting regional differences in mortality? Addressing these questions of how mortality differ across districts whilst also how this is linked to regionally measurable variables, this study now looks at the effect and role of individuals and districts levels characteristics on regional mortality variations among older adults in India. It is apparent that regional affiliations are influenced by the attributes of persons residing across various parts of India. As a consequence, policymakers must take into account regional variations in mortality in order to build more relevant and targeted programs and interventions.

Therefore, we intend to evaluate the effect of individuals and districts levels determinants on regional mortality variations among older adults in India. Additionally, we have performed decomposition analysis to evaluate the role of socioeconomic and demographic dimensions within regional mortality variations among older adults in India.

### 1.1 Some International evidence

Mortality measures at the national level often hide the essential within-regional variations in a country. According to a study, those who reside in regions with poor socioeconomic conditions have a greater rate of mortality (Meijer et al., 2012). Earlier study has identified substantial cross-level interactions (across all outcome measures), implying that the effect of individual-level variables on mortality measures changes by context (Riva et al., 2007). Previous research has claimed of changing mortality regime identified in most Western and Central provinces of Canada among women, although all men remained to experience mortality compression regime among the older populations. (Ouellette et al., 2013). In comparison to the impacts of individual socioeconomic attributes, studies in numerous countries like-United States (Steenland, 2004), Helsinki (Martikainen, 2003) and Germany (Kibele, 2014) have revealed that the effects of socioeconomic factors of area on mortality are relatively small. Changes in the socioeconomic and demographic factors have influenced the areas in different ways, resulting in increasing regional differences and shifts in population distribution (Bontje & Musterd, 2012; Martinez-Fernandez et al., 2012; Suulamo et al., 2021). Recent study has showed significant geographical differences in mortality fall among older people in China (Zhang et al., 2022). Studies have documented that even the mortality gaps are reduced through the time in some countries; others have persisted and even widened as the general mortality rate has decreased. (Suulamo et al., 2021; Vierboom et al., 2019; Wilson et al., 2020).

## 2. Data and methods

### 2.1 Data source

The present study has used the 4th round of Demographic and Health Survey (DHS) conducted in 2015-2016 in India by the Ministry of Health and Family Welfare (Government of India). It is often referred as National Family Health Survey-4 (NFHS-4) (IIPS & ICF International, 2017). It covers information on fertility, maternal and reproductive health, women’s and children’s nutritional status, infant, family planning, the quality of health services, child morbidity and mortality at the national, state, and district levels (IIPS & ICF International, 2017). There are 425,563 rural households and 175,946 urban households in the study. A total of 699,686 women aged 15 to 49 were interviewed, with a 97 percent response rate. The NFHS-4 sample size was chosen to generate indicators at the states, union territories (UTs) and district levels. The sample was chosen in two stages: rural with villages as Primary Sampling Units (PSUs) at the first stage (selected with probability proportional to size), and urban with Census Enumeration Blocks (CEB) followed by a random selection of 22 households in each PSU and each CEB, respectively, at the second stage (selected with probability proportional to size). After executing a full mapping and household listing operation in the designated first-stage units in both urban and rural areas, households were selected for the second stage.

### 2.2 Outcome variable

Since January 2013, the NFHS-4 has been compiling data on mortality in the ‘household.’ The survey collected information on the deceased persons’ age at death, sex, month, and year of death if any deaths happened between January 2013 and the survey. Thus, the outcome variable in the study is the mortality among older adults (or 60+ individuals) and it is dichotomous and categorized into two categories where ‘1’= deceased persons (60+ individuals) and ‘0’= alive persons (60+ individuals). However, our analysis is based on individual level data and district level data.

Our study has categorized the regions into six regional zones are as follow:

1. Northern (Jammu and Kashmir, Delhi, Haryana, Himachal Pradesh, Chandigarh, Punjab, Uttarakhand, and Rajasthan)
2. North-eastern (Assam, Arunachal Pradesh, Meghalaya, Manipur, Nagaland, Mizoram, Tripura and Sikkim)
3. Central (Madhya Pradesh, Uttar Pradesh, and Chhattisgarh)
4. Eastern (Odisha, Bihar, Jharkhand, West Bengal)
5. Western (Goa, Maharashtra, Gujarat, Daman & Diu, and Dadra & Nagar Haveli)
6. Southern (Tamil Nadu, Kerala, Telangana, Andhra Pradesh, Karnataka, Puducherry, Andaman and Nicobar Islands, and Lakshadweep)

### 2.3 Exposure variables

#### 2.3.1 Individual-level determinants

Age-groups is categorized into three such as young old which is 60-69 years age groups, middle-old is 70-79 years, oldest-old is 80+ years. Sex is classified into two such as male and female while Place is categorized into two such as rural and urban. We have also used caste groups which has three groups such as Schedule caste (SC) & Schedule Tribe (ST), Other backward caste (OBC) and general (no caste). In case of religion, it has three main categories such as Hindus, Muslims and others including Christians, Buddhist, Sikhs, Jews, Parsis etc. Our study has also take income group which is categorized into five, such as Poorest, Poorer, Middle, Richer and Richest respectively.

#### 2.3.2 District-level determinants

Based on the district level characteristics, our study has included five important characteristics in the analysis: a) Proportion of 60+ people in the district, b) Proportion of literate population in the district, c) Proportion of district level health insurance coverage, d) District level proportion of electricity, e)District level proportion of public health facilities. The region of residence has been applied with the following geographical categorizations with six major regions mentioned above. But besides that all other variables at district-level are established by combining the individual attributes within their cluster, with the exception of the regions variable. The proportions’ average of the individual in each of the categories of a particular factor is used to compute these aggregates for clusters respectively.

### 2.4 Statistical analysis

Our study has carried out univariate, bivariate and multilevel analyses. We have analysed descriptive at univariate level in order to investigate the respondents’ sample distribution. We performed cross-tabulation at bivariate level and we also carried out the chi square test which was developed by Pearson to investigate the relationship between the outcome variable and the other specified exposure variables. And then we have performed a two-level logistic regression model to look into the impact of individual and district levels determinants of mortality among older adults and to determine the scope by which they elucidate the regional mortality variations among older adults. Additionally, we have performed decomposition analysis to evaluate the contribution of socio-economic and demographic determinants within regional mortality variations among older adults in India. The decomposition approach used in the study was proposed by (Powers et al., 2011). The data were analysed using R (version) for multilevel and STATA 14.0 for decomposition analysis.

### 2.5 Multi-level logistic model

Two level model (individual level and district level)

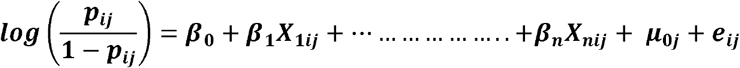

Where, 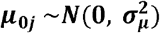

Where ***p***_***ij***_ is the risk of old-age mortalityAlso, elderly death is the binary response variable for individual ***‘i’*** and distrcit ***‘j’***. ***β***_**0**_ intercept measuring the log of odd ***β***_**1**_ **… … … … … *β***_**n**_ are size effect of individual and district-levels determinants, ***X***_**1*ij***_ **… … … … … *X***_***nij***_ are exposure vaeiables at individuals and districts-levels.

### 2.6 Decomposition approach

Our study has asopted the multivariate decomposition approach which was proposed by Powers et al 2011. This approach is based on non-linear response outcomes where we have tested the inter-regional variations in mortality among older adults. We carried separate decompositions where Northern-region has been considered as a low outcome group while the high outcome groups are North-eastern, Central, Eastern, Western and Southern regions, respectively. The total disparity in a measured result can be decomposed into a sum of components due to group variations in risk variables and group differences in the influence of those dimensions (Powers, 2016). However, the gaps in total rates between two regional groups, ‘***a***’ and ‘***b***’, would be subdivided or decomposed as follows:

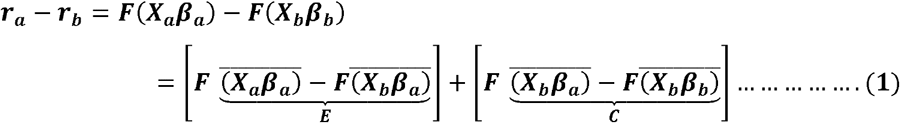

Where 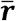 is denoted as the overall mortality among older adults in each population of the respective region and ***F***(***X***_***β***_) is denoted as a linear combination of risk factors which is mapped by a differentialble function X and effect ***β*** in the multivariate model described below:

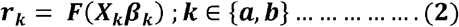

From the above equation **(2)**, ‘***r***’ is denoted as ***N*** × **1** rate vectors, **X** is ***N*** × ***P*** matrix of the exposure variables, and ‘***β***’ is a ***P*** × **1** coefficients of logistic regression in a vector. The multivariate model’s findings are derived independently for each variable across regions. We have selected the Northern region as a reference group (the group labelled as ‘b’) while the comparative group is either the North-eastern, Central, Eastern, Western or Southern regions (the group labelled as ‘a’). In the preceeding equation (1), the total gaps in the mortality among older adults are classified into two parts using the multivariate decomposition: endowment (E) and coefficient (C). The “endowment” is the portion of a change in the composition of a collection of indicators while the “coefficient” is the portion of the change in the influence of the indicators in the analysis that can be attributed to the change in the coefficient (Bora, 2020). For example —if we take mortality among older adults as a outcome of interest and districts covariates as a determinant affecting mortality among older adults. We then decompose the overall changes in mortality among older adults due to district covariates into ‘endowment’ and ‘coefficient’ componnets, then the ‘endowment’ component is the contrbuted by the change in the district covariates, and the ‘coefficient’ component is contributed by influencing the district covariates on mortality among older adults (Bora, 2020). Likewise, our decomposition technique has addressed the key questions challenging the potential implications of equalizing attributes across the groups or regions, respectively.

## 3. Results

### 3.1 Individual level characteristics by regions

Table 1 presents the percentage of the sample distributions of the individual characteristics among older adults in India and regions with suitable background characteristics using DHS data 2015-16. Eastern region is showing the highest percentage of the young-old population with 64.58% while the lowest is seen in North-eastern region with 60.63%. The older male percentage is found to be greatest in North-eastern region (53.73%) whereas western region is showing the lowest percentage with 48.49%. Likewise, the highest urban population is observed in Southern region with 35.29% and lowest in Eastern region (17.02%). Interestingly, the percentage of SC/ST is highest in North-eastern region (68.05%), OBC are highest in Southern region with 58.52% and general (no caste) is highest seen in Northern region. Central region is showing the highest percentage of Hindus (87.94%) and Muslims (10.9%). However, Eastern region is showing the highest percentage of poorest older indivudals while the highest richest is observed in Northern region.

**Table 1.** Percentage distribution of individual level characteristics for older adults by regions in India, DHS (2015-16).

| **Background characteristics** | **India** | **Northern** | **North-eastern** | **Central** | **Eastern** | **Western** | **Southern** |
| --- | --- | --- | --- | --- | --- | --- | --- |
| **Age-groups** |  |  |  |  |  |  |  |
| Young-old | 62.78 | 62.56 | 60.63 | 61.82 | 64.58 | 62.65 | 64.08 |
| Middle-old | 26.37 | 26.13 | 26.9 | 27.07 | 25.12 | 27.16 | 26.21 |
| Oldest-old | 10.85 | 11.31 | 12.48 | 11.11 | 10.3 | 10.19 | 9.71 |
| **Sex** |  |  |  |  |  |  |  |
| Male | 51.26 | 51.43 | 53.73 | 50.93 | 52.9 | 48.49 | 49.55 |
| Female | 48.74 | 48.57 | 46.27 | 49.07 | 47.1 | 51.51 | 50.45 |
| **Place** |  |  |  |  |  |  |  |
| Rural | 73.97 | 72.65 | 74.8 | 75.57 | 82.98 | 68.22 | 64.71 |
| Urban | 26.03 | 27.35 | 25.2 | 24.43 | 17.02 | 31.78 | 35.29 |
| **Caste** |  |  |  |  |  |  |  |
| SC/ST | 33.56 | 29.6 | 68.05 | 29.41 | 33.32 | 28.86 | 24.54 |
| OBC | 40.79 | 29.75 | 14.78 | 49.69 | 45.31 | 34.74 | 58.52 |
| General | 25.65 | 40.64 | 17.17 | 20.91 | 21.36 | 36.4 | 16.94 |
| **Religion** |  |  |  |  |  |  |  |
| Hindu | 78.55 | 75.45 | 38.61 | 87.94 | 85.77 | 85.23 | 82.26 |
| Muslims | 8.96 | 8.75 | 4.41 | 10.9 | 9.41 | 7.44 | 9.75 |
| Others | 12.48 | 15.8 | 56.98 | 1.15 | 4.82 | 7.33 | 7.99 |
| **Income** |  |  |  |  |  |  |  |
| Poorest | 21.3 | 8.33 | 13.01 | 31.58 | 41.58 | 13.19 | 7.43 |
| Poorer | 20.26 | 15.09 | 28.33 | 21.49 | 23.29 | 18.2 | 17.09 |
| Middle | 19.46 | 18.08 | 27 | 15.87 | 16.04 | 21.25 | 25.24 |
| Richer | 18.38 | 20.78 | 19.51 | 13.94 | 11.39 | 23.18 | 27.52 |
| Richest | 20.6 | 37.73 | 12.15 | 17.12 | 7.7 | 24.18 | 22.72 |
| **Total (%)** | **100** | **20.7** | **10.71** | **25.03** | **18.93** | **9.96** | **14.63** |
| **Total (n)** | **2,71,679** | **56,342** | **29,105** | **67,995** | **51,433** | **27,055** | **39,749** |

Source: Authors' own calculation using Demographic Health Survey data (2015-16).

Table 2 presents the percentage distribution of mortality among older adults by regions in India with suitable background characteristics using DHS data (2015-16). Our result shows that the highest share of the mortality among older adults comes from Cental region (12.28%) followed by Eastern region (11.31%) and North-eastern (10.81%) while the lowest share is observed in Northern region (9.24%) followed by Southern region (9.28%). We found that the risk factors for mortality among older adults are subtantial higher in the Central, Eastern and North-eastern regions of India such as age-groups, sex, place of residence, caste groups, religion groups, and income groups respectively.

**Table 2.**
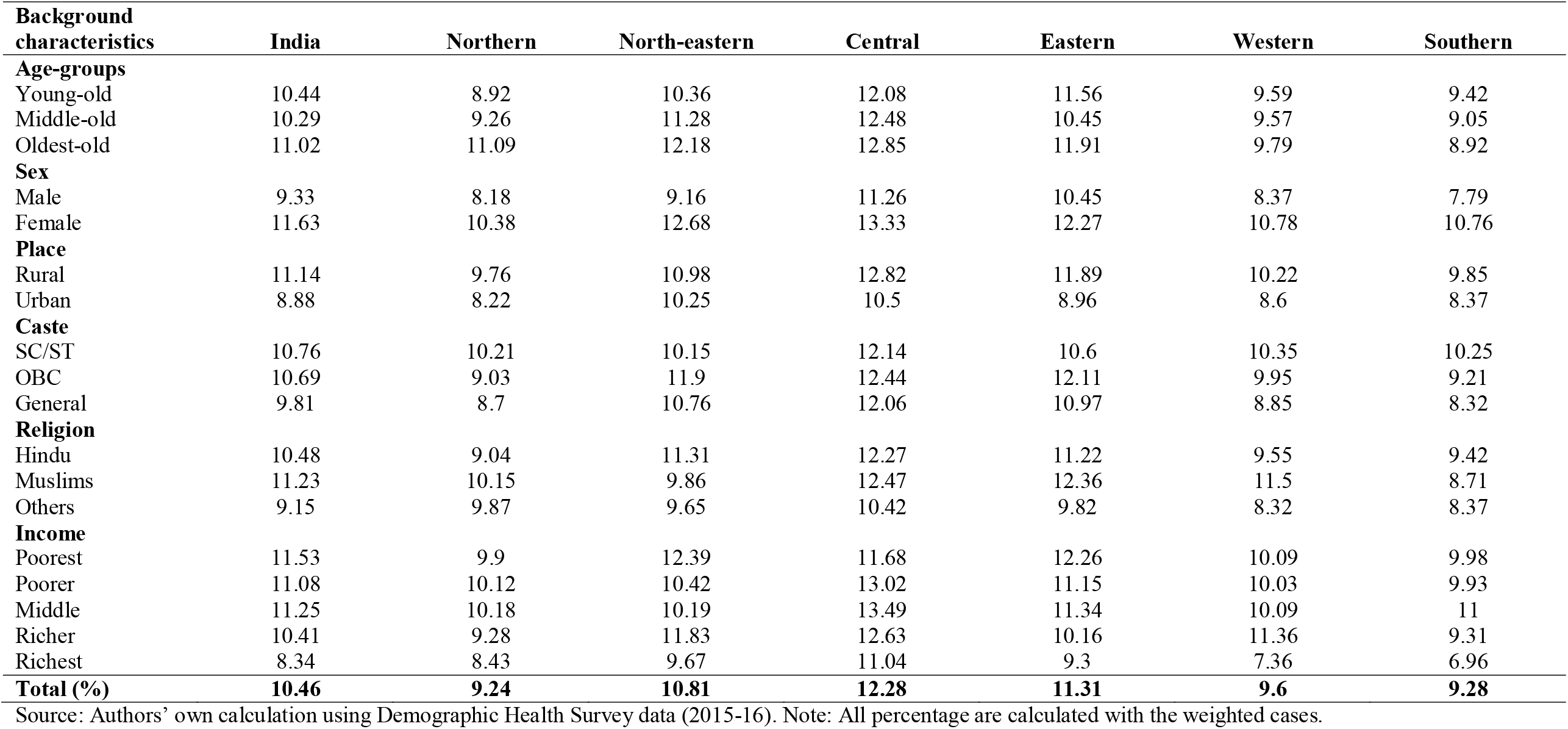
Percentage distribution of mortality among older adults by regions in India with background characteristics, DHS (2015-16).

| **Background characteristics** | **India** | **Northern** | **North-eastern** | **Central** | **Eastern** | **Western** | **Southern** |
| --- | --- | --- | --- | --- | --- | --- | --- |
| **Age-groups** |  |  |  |  |  |  |  |
| Young-old | 10.44 | 8.92 | 10.36 | 12.08 | 11.56 | 9.59 | 9.42 |
| Middle-old | 10.29 | 9.26 | 11.28 | 12.48 | 10.45 | 9.57 | 9.05 |
| Oldest-old | 11.02 | 11.09 | 12.18 | 12.85 | 11.91 | 9.79 | 8.92 |
| **Sex** |  |  |  |  |  |  |  |
| Male | 9.33 | 8.18 | 9.16 | 11.26 | 10.45 | 8.37 | 7.79 |
| Female | 11.63 | 10.38 | 12.68 | 13.33 | 12.27 | 10.78 | 10.76 |
| **Place** |  |  |  |  |  |  |  |
| Rural | 11.14 | 9.76 | 10.98 | 12.82 | 11.89 | 10.22 | 9.85 |
| Urban | 8.88 | 8.22 | 10.25 | 10.5 | 8.96 | 8.6 | 8.37 |
| **Caste** |  |  |  |  |  |  |  |
| SC/ST | 10.76 | 10.21 | 10.15 | 12.14 | 10.6 | 10.35 | 10.25 |
| OBC | 10.69 | 9.03 | 11.9 | 12.44 | 12.11 | 9.95 | 9.21 |
| General | 9.81 | 8.7 | 10.76 | 12.06 | 10.97 | 8.85 | 8.32 |
| **Religion** |  |  |  |  |  |  |  |
| Hindu | 10.48 | 9.04 | 11.31 | 12.27 | 11.22 | 9.55 | 9.42 |
| Muslims | 11.23 | 10.15 | 9.86 | 12.47 | 12.36 | 11.5 | 8.71 |
| Others | 9.15 | 9.87 | 9.65 | 10.42 | 9.82 | 8.32 | 8.37 |
| **Income** |  |  |  |  |  |  |  |
| Poorest | 11.53 | 9.9 | 12.39 | 11.68 | 12.26 | 10.09 | 9.98 |
| Poorer | 11.08 | 10.12 | 10.42 | 13.02 | 11.15 | 10.03 | 9.93 |
| Middle | 11.25 | 10.18 | 10.19 | 13.49 | 11.34 | 10.09 | 11 |
| Richer | 10.41 | 9.28 | 11.83 | 12.63 | 10.16 | 11.36 | 9.31 |
| Richest | 8.34 | 8.43 | 9.67 | 11.04 | 9.3 | 7.36 | 6.96 |
| **Total (%)** | **10.46** | **9.24** | **10.81** | **12.28** | **11.31** | **9.6** | **9.28** |

Source: Authors' own calculation using Demographic Health Survey data (2015-16). Note: All percentage are calculated with the weighted cases.

### 3.2 District level characteristics by regions

Table 3 presents the descriptive statistics of the district level characteristics by regions in India. Most of the older individuals (60 years and above) residing in the districts are found in all regions. Considering the older individuals residing in the districts of North-eastern region where the mean of literate population are showing lower. The average health insurance coverage among the older people is found to be lower in the districts of Central region. Older individuals living in the districts of North-eastern region followed by Central region have lower access to electricity. On the other hand, older individuals residing in the districts of Central region is showing lower access of public health facilties or Government hospitals.

**Table 3.** Descriptive statistics of district-level characteristics by regions in India, DHS (2015-16).

| District level variables | Mean (Std. Deviations) |  |  |  |  |  |  |
| --- | --- | --- | --- | --- | --- | --- | --- |
|  | India | Northern | North-eastern | Central | Eastern | Western | Southern |
| Proportion of 60+ people in the district | 8.4(2.06) | 8.37(2.2) | 8.32(1.8) | 8.72(1.71) | 8.51(2.1) | 7.36(2.34) | 8.48(2.1) |
| Proportion of literate population in the district | 0.63(0.1) | 0.62(0.1) | 0.61(0.11) | 0.64(0.08) | 0.62(0.12) | 0.67(0.09) | 0.62(0.11) |
| Proportion of district level health insurance coverage | 0.27(0.22) | 0.37(0.27) | 0.34(0.22) | 0.17(0.12) | 0.26(0.18) | 0.19(0.18) | 0.3(0.26) |
| District level proportion of electricity supply | 0.88(0.15) | 0.93(0.11) | 0.8(0.15) | 0.85(0.16) | 0.89(0.16) | 0.96(0.06) | 0.88(0.14) |
| Proportion of marginal workers in the district | 0.11(0.06) | 0.11(0.06) | 0.14(0.06) | 0.1(0.05) | 0.09(0.05) | 0.11(0.06) | 0.11(0.04) |
| Proportion owned-house in the district | 0.8(0.14) | 0.79(0.15) | 0.83(0.11) | 0.83(0.13) | 0.77(0.14) | 0.75(0.16) | 0.84(0.11) |
| Proportion of urban population in the district | 0.27(0.22) | 0.29(0.22) | 0.18(0.18) | 0.26(0.19) | 0.33(0.27) | 0.26(0.19) | 0.26(0.18) |
| District level proportion of public health facilities | 0.53(0.25) | 0.66(0.23) | 0.68(0.3) | 0.4(0.18) | 0.49(0.2) | 0.67(0.23) | 0.45(0.23) |

Source: Authors' own calculation using Demographic Health Survey data (2015-16).

### 3.3 Risk factors of mortality among older adults by regions: Multilevel Analysis

Table 4 presents the results of multilevel analysis. The first model (Null model or Model ‘0’), which has no explanatory variables and clearly indicates significant mortality variations among older adults which is explained between the cluster variation of the characteristics (ICC=0.75%, p<0.01). The Model 1 is showing only the region covariate, in order to see if the mortality risk among older adults differed by regions. The result illustrates that the older individual residing in the Central (OR=1.27;p<0.01) and Eastern (OR=1.15; p<0.1) regions are havingh greater mortality risk compared to Norther Region of India. The covariate of regions is included in the multilevel model to see the significant variation across individual and district levels. The Model 2 is showing the sociodemographic and economic covariates of the older individuals. The North-eastern (OR=1.02; p<0.01) and Central (OR=1.24; p<0.1) regions are showing greater mortality risks among older individuals compared to Northern region.

**Table 4.**
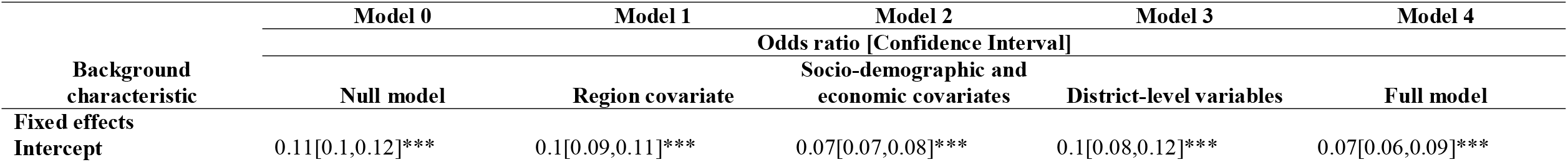
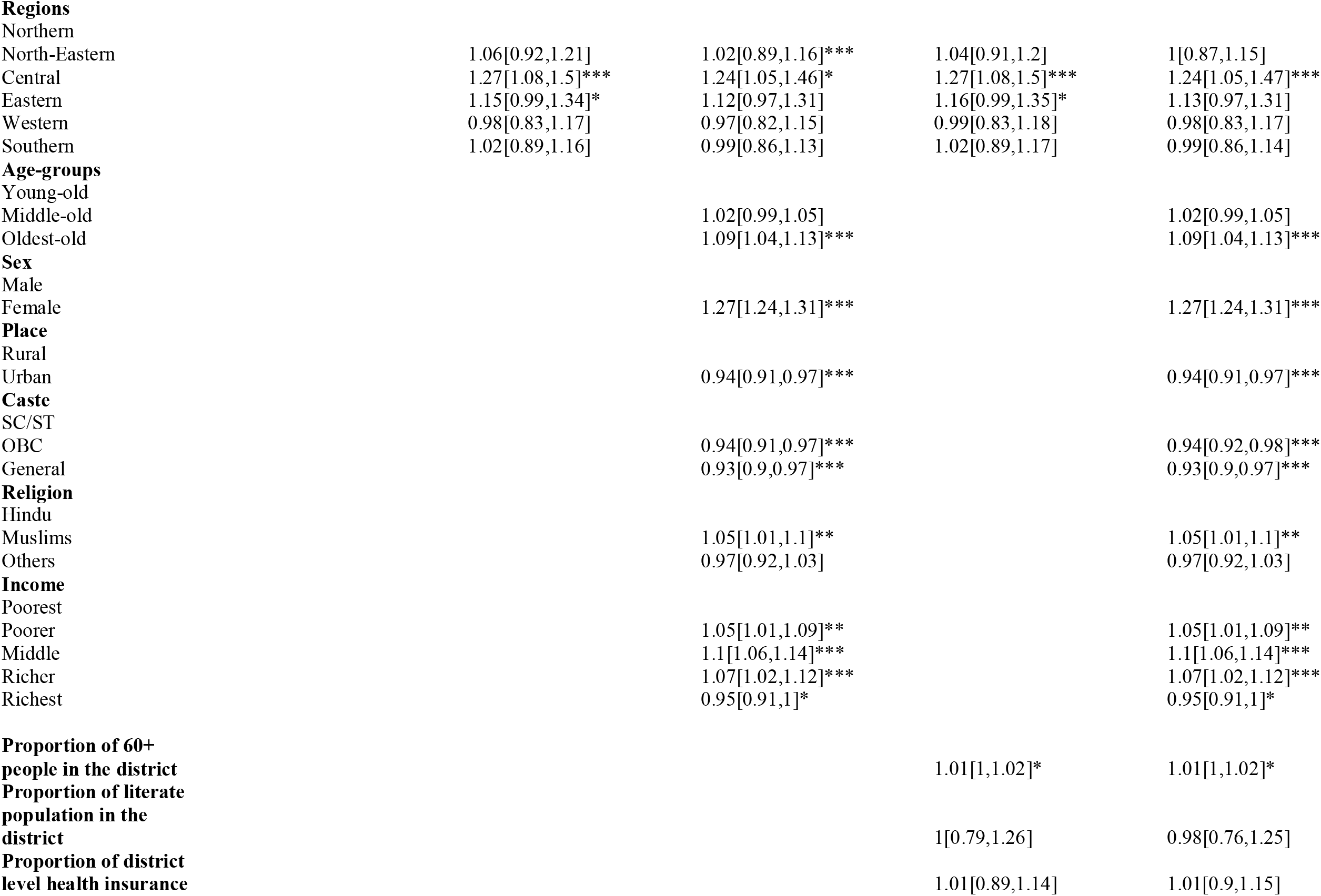
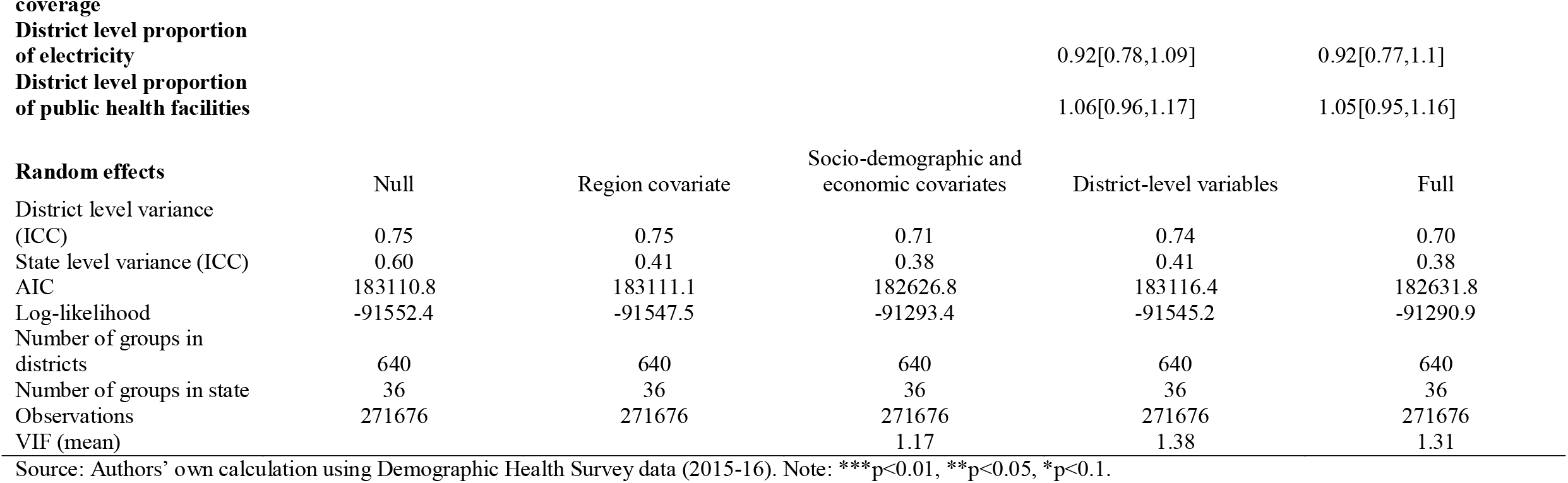
Multilevel logistics regression model showing factors associated with mortality among older adults in India, DHS (2015-16).

However, the selected district-level covariates are considered in the Model 3. The results showed that when just district-level covariates were included in the multilevel models, the mortality risk in regions is remained higher in Central (OR=1.27; p<0.01) and Eastern (OR=1.16; p<0.1) regions compared to Northern regions. The full model (Model 4) which represents the individual- and district-level characteristics including the regions, district-level 60+ populations, literate population in the district, district-level health iinsurance coverage, district-level access to electricity, district-level public health facilities, as well as individual-level characteristics such as sex, place of residence, caste-group, religions, household income that are significant predictors of mortality among older adults in India. For instance- the result from the Model 4 indicating that the Central region (OR=1.24; p<0.01) is showing significantly greater mortality risk among older adults compared with Norther region. The oldest-old age group (OR=1.09; p<0.01), older females (OR=1.27; p<0.01) and Muslims (OR=1.05; p<0.05) are showing higher mortality risks among older adults. However, the older individuals belonging to the household income group pooerest (OR=1.05; p<0.05), Poorer (OR=1.1; p<0.01) and Richer (OR=1.07; p<0.01) are observed at greater mortality risk while the Richest (OR=0.95; p<0.1) income group showing the lower mortality risk. The 60+ people in the distrct is also showing greater mortality risks with (OR=1.01; p<0.1).

### 3.4 Findings from the decomposition analysis

Table 5 presents the multivariate decomposition of regional mortality variations showing the contribution of mortality gap among the older adults, which are contributed to the gaps in endowment (E) and to gaps in coefficients (C) in India using DHS data (2015-16). The results assess each risk factor that contributes to each component of mortality differences among older adults across the regions. Our finding shows that the “Northern regions versus North-eastern regions” column reflect the elements of the Northern-North-eastern regional variations in mortality among older adults, considering the substantial contributions to the mortality differences that are illustrated in columns E (endowment) and C (coefficient) (district). The following is an interpretation of these elements: If the risk factor distribution in the Northern region remains unchanged, however, in the north-eastern region, those risk factors had no influence., a negative (positive) E coefficient implies a gain (loss) or increase (decrease) in the mortality difference among older adults. Similarly, if there is no change in the effect of risk factors in the Northern region, the effect of risk factors in the North-eastern region is unaffected. Thus, a negative (positive) C coefficient indicates a gain (loss) or increase (decrease) in the mortality gaps. Likewise, the remaining regional group variations can be interpreted similarly.

**Table 5.**
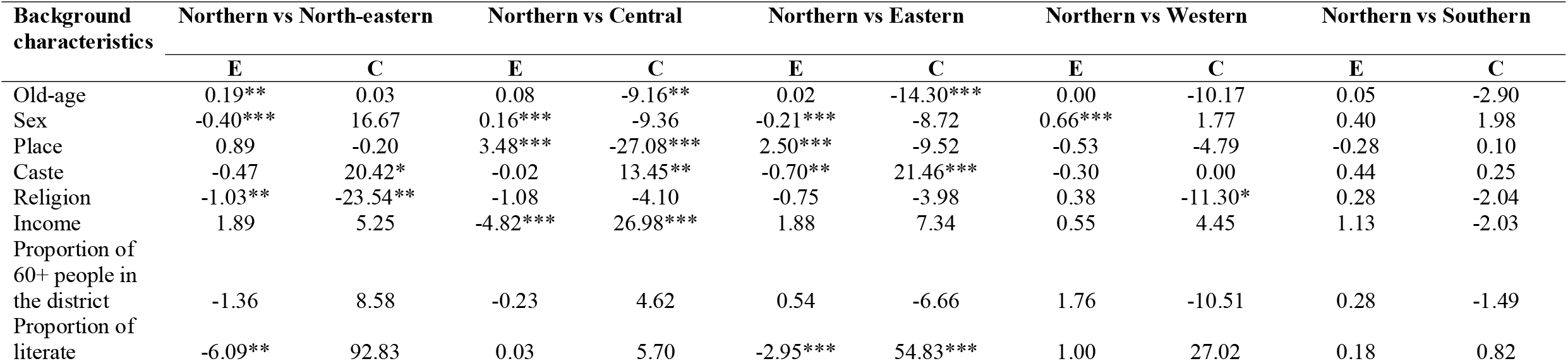
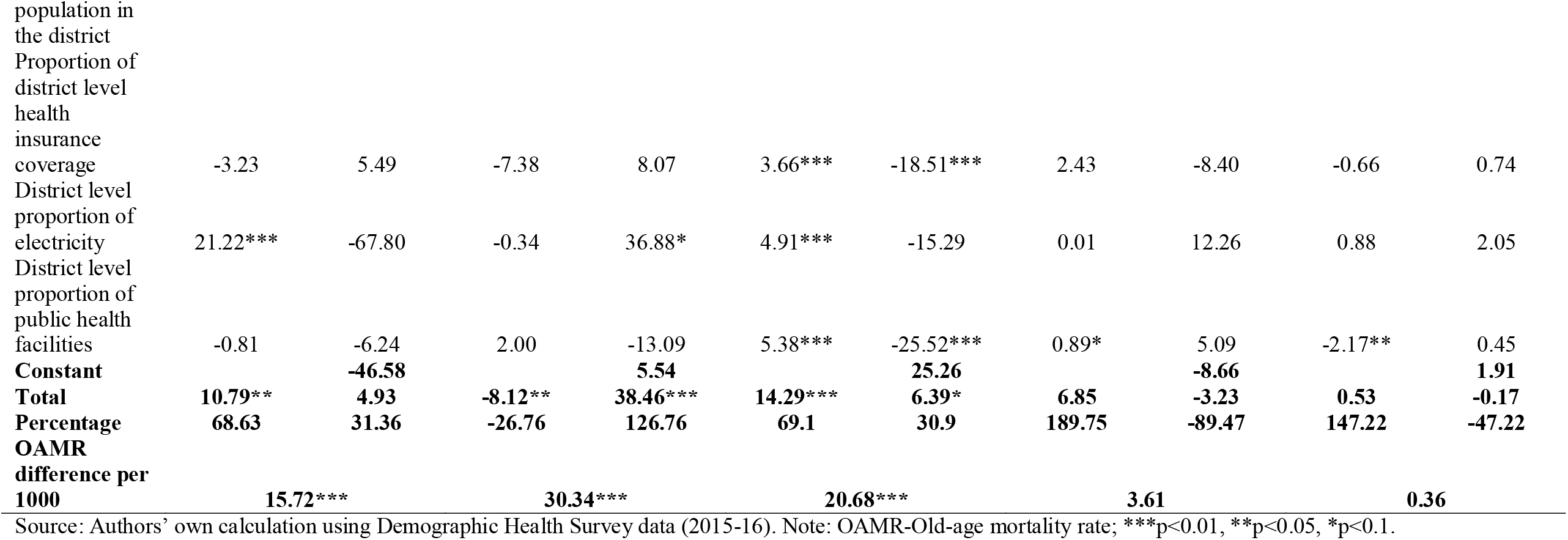
Multivariate decomposition of regional difference in old-age mortality showing contribution to old-age mortality gap attributed to differences in endowment (E) and to differences in coefficient (C) in India, DHS (2015-16).

The decomposition results between the Northern and North-eastern regions indicate the endowment (E) gaps explaining 68.63% of the observed regional mortality variations among older adults. The endowment differences accounted for -26.76% of the given mortality gaps between the Northern and the Central regions. Similarly, the differences in endowment between Northern and Eastern, Northern and Western, and Northern and Southern regions contribute around 69.1%, 189.75%, and 147.22% of the difference in mortality among older adults. However, the Central region is indicating significant higher mortality gaps compared to the Northern region, which is 30 deaths per 1000 individuals, while the Eastern region (20.68 deaths per 1000 individuals) and North-eastern region (15 deaths per 1000 individuals) are also showing significant mortality gaps among older adults compared to Northern region.

Considering the comparison between Northern and North-eastern regions, we found that there is a positive contribution of the endowment (E) effect, which are significantly associated with the old-age group variable (0.19; p<0.05) and access to electricity in the district variable (21.22; p<0.01), if the effect for North-eastern region is unchanged while comparing to other regional groups, then OAM gap is expected to decrease by 21 deaths per 1000 individuals. Sex (−0.4; p<0.01), religion (−1.03; p<0.05), the literate population in the district (−6.09; p<0.05) variables are showing significantly negative contributions of the endowment (E) to the OAM gaps, if the effect is fixed then the OAM difference is expected to increase by 7 deaths per 1000 individuals. On the other hand, we found that the Caste group (20.42; p>0.1) shows a positive contribution to the coefficient (C) effect to the OAM gaps while religion (−23.54; p<0.05) indicates a negative contribution.

Likewise, comparing between Northern and Central regions, our finding indicates a positive contribution of the E effect, which is significantly associated with sex (0.16; p<0.01) and place (3.48; p<0.01). If the effect in Central regions remains the same, the OAM gaps are expected to decrease by 3 deaths per 1000 individuals. While the income (−4.82; p<0.01) is significantly negatively contributing to the endowment effect in OAM gaps, where the OAM gaps are expected to increase by 4 death per 1000 individuals. Besides that, old-age (−9.16; p<0.05) and place (−27.08; p<0.01) are negatively contributing to the C effect in OAM gaps, indicating that if the Central regional effect remains the same, then it is expected to increase by 36 deaths per 1000 individuals. However, the caste group, income group, access to electricity positively contribute to the C effect in OAM gaps.

Furthermore, we compare the regions between Northern and Eastern zones; the finding shows a positive contribution of the E effect which is significantly associated with the place (2.5; p<0.01), insurance coverage at district level (3.66; p<0.01), access to electricity at district level (4.91; p<0.01), access to public health facilities (5.38; p<0.01). If the effect in the Eastern regions is unchanged, then the OAM gaps are expected to decrease by 16 deaths per 1000 individuals. Whereas the sex (−0.21; p<0.01), caste (−0.7; p<0.05), and the literate population at district level (−2.95; p<0.01) are significantly negatively associated with the endowment effect in OAM gaps, such that OAM gaps may increase by 3 deaths per 1000 individuals if the effect in the Eastern region will remain same. On the other hand, old-age groups (−14.3; p<0.01), insurance coverage at district level (−18.51; p<0.01), access to public health facilities (−25.52; p<0.01) are significantly negatively contributing to the C effect in OAM gaps, where the OAM gaps may increase by 58 deaths per 1000 individuals if the effect in Eastern region showing no change. While the caste group (21.46; p<0.01) and the literate population at the district level (54.83; p<0.05) are showing a positive contribution to the C effect in OAM.

Meanwhile, if we compare the regions between Northern and Western, our finding indicates a positive contribution to the E effect, which is significantly associated with sex (0.66; p<0.01) and access to public health facilities (0.89; p<0.1). If the effect in the Western region remains unchanged, then the OAM gaps are expected to decrease by 1 death per 1000 individuals, which is small. Religion (−11.3; p<0.1) is negatively contributing to the C effect in OAM gaps, where the OAM gaps may increase by 11 deaths per 1000 individuals if the effect in the Western region remains fixed.

Nevertheless, lastly, we compare the regions between Northern and Southern. We found a negative contribution in the E effect, which is significantly associated with access to public health facilities (−2.17; p<0.05). If the effect in the Southern region remains unchanged, then OAM gaps are expected to increase by 2 deaths per 1000 individuals.

## 4. Discussion

Our study has investigated the impact of individual and district-level determinants on mortality among older adults in India and we have also assessed the extent how they impacted the regional mortality variation among older adults in India using the Demographic Health Survey (DHS, 2015-16). Our finding revealed that mortality risk among older adults in India is impacted by both individual and district-level determinants. Our finding has quantified the determinants that contribute to the regional mortality variations among older adults in India.

The multilevel analyses results revealed that regional mortality variations exist at both individual and district levels among older adults in India. Our finding identified that older adults living in the Central region have a higher mortality risk than in Northern regions. There are some plausible explanations for these regional differences. The greater mortality risk is observed in the Central region that could be due to socio-demographic and economic reasons. Though Central region has a shorter life expectancy and a smaller proportion of older individuals than the Northern region (SRS Report, 2019). In the Central region, poor health-care facilities and negligence of visiting health care centres are also linked to a higher death risk among older adults (Akhtar & Saikia, 2022; Banerjee, 2021). As a result, it is recommended that each state in the central region emphasize the importance of disease prevention and control, raise disease prevention awareness, and develop strategic scientific measures to reduce mortality risks and control critical areas and populations while taking into account the current situation in the region.

However, our finding revealed that oldest-old has greater mortality risk compared to young-old and similar results are also reflected in Australia (Khalatbari-Soltani et al., 2020), France (Menvielle et al., 2010) and in other European countries (Huisman, 2004; Huisman et al., 2013). Meanwhile, our study has also identified that older females have higher mortality risk compared to older males. The plausible reason could also be the lower educational status among females as evident in the previous study (Mackenbach et al., 1999; Rostad et al., 2009). While greater female mortality is also observed in Finland and Spain but contrasting result is seen in Israel and Netherland where older male mortality is at greater risk (Noale et al., 2005). However, our study has also revealed that urban residence has lower mortality risk among older adults while consistent finding are seen in Germany and England & Wales (Ebeling et al., 2022) while China (Zhao et al., 2020) showed a contradictory result from our findings. The different distribution in the access to health care facilities and health care spending between rural and urban place of residence could potentially be a contributing factor (Saikia et al., 2013).

Furthermore, ur study has found that older adults belonging to poorer, middle, and richer household income groups have significantly greater mortality risks but lower among richest compared to poorest. Though earlier studies evident a positive association between income and mortality among older adults (Huisman, 2004; Huisman et al., 2013; Rehnberg, 2019). Additionally, our study has revealed a significantly lower mortality risk among older adults belonging to the General and OBC caste compared to SC/ST caste but a higher mortality risk among Muslims than Hindus. Though SC/ST and Muslims are considered as marginalized social groups in India and they are socio-economically poorer (Desai, 2010; Thorat & Newman, 2010; Vyas et al., 2022), they also have lower life expectancies (Kumari & Mohanty, 2020).

Lastly, our decomposition analysis results revealed a significant regional mortality variation among Indian older adults, where the Central region has the significantly greater average number of excess mortality, followed by Eastern and North-eastern regions compared to other regions. However, the gaps in the overall regional mortality among older dults is relatively higher between Northern-Central and Northern-NorthEastern. Interestingly, old-age, sex, place of residence, caste, religion and household income have showed significant determinants that contribute to the regional mortality variation among older adults in India. Desptite that district-level literacy, insurance coverage, electricity supply and public health facilities showed a significant impact on district level mortality among older adults in India. It is suggested that several social factors-including education, health insurance and better health care facilities could shape and improve the health care attitude and behaviour. On the other hand, awareness and improvement in treatment seeking behaviour could also result in reducing mortality risks among older adults.

## 5. Strength of the study

Our research is based on nationally representative cross-sectional datasets from the demographic health survey (2015-16). Hence, our findings are clearly applicable to the country’s health policy implementation. Our findings clearly enable us to focus, design and implement the health interventions among the older people in India at the district level. This will be helpful for the stakeholders, policymakers and health workers in order to improve the health conditions, health institutions and wellbeing of older people in India. Furthermore, it will also be helpful to avert the deaths among older adults in India, which directly attains Goal 3 of SDG by ensuring healthy lives and promoting wellbeing for all people.

## 6. Limitations

Despite a comprehensive analyses, our study has some limitations. First, with the exception of the place of residence and region variables, additional district-level variables are generated by combining the data with individual-level characteristics at the cluster level. There is a possibility that autocorrelation has occurred while preparing the district-level variables. As a result, we performed a correlation test in order to mitigate the problem in the data. Henceforth, several essential district-level characteristics such as proximity to the educational level of the deceased person, cause of death by disease, marital status, living arrangements and other factors such as environmental, life style and behavioral factors could not be included in the study because of the data unavailability. Lastly, a selective in-and-out migration is also of particular concern when comparing regional mortality variation. The health state of migrants and the reasons to relocate, which differ at older ages, determine whether old-age mortality falls or rises as a result of migration. Such information was not available in the survey data. Therefore, we could not include.

## 7. Conclusions

In overall, the findings of the present study are extremely important for India’s public health. We intended to evaluate the effect of individuals and districts levels determinants on regional mortality variations among older adults in India. Additionally, we have performed decomposition analysis to evaluate the role of socioeconomic and demographic dimensions within regional mortality variations among older adults in India using the DHS datasets 2015-16. Our finding has revealed that the risk of mortality among older adults is mainly contributed by individual and district level characteristics. Our study has found the significant determinants of the regional mortality variations among older adults in India. Our results showed that the regional mortality variations are remained higher in the Central regions even after including the district-level covariates in the multilevel analyses.

Our findings from the decomposition analysis revealed that the Central, Eastern and North-eastern regions have the higher average number of excess death compared to other regions. Hence, there is a need to reduce these mortality disparities by improving the health facilities at both public and private settings, social services, awareness of public services at the regional level. In order to eliminate these mortality gaps, there is a need for solid support from the state and central government to bridge the socio-demographic and economic development in India at the regional level. As a result, policy should include efforts to improve health outcomes among older adults at early stages.

## Data Availability

The datasets are publicly available and can be accessed using the link https://dhsprogram.com/data/available-datasets.cfm

